# Impacts of regional climate on the COVID-19 fatality in 88 countries

**DOI:** 10.1101/2020.06.13.20130013

**Authors:** Minneng Wen, Liyuan Chen

## Abstract

The COVID-19 pandemic had led to 500000 confirmed death by June 30, 2020. We combined the number of monthly confirmed new cases and deaths with latitude, temperature, humidity, rainfall, and sunshine ultraviolet (UV) to explore the climate impacts on COVID-19 fatality in 88 countries. There was a significant decrease in overall case-fatality rate in May and June (from 8.17% to 4.99% and 3.22%). The fatality in temperate marine regions was the highest (11.13%). The fatality was 5.71% in high latitudes (≥30°) but only 3.73% in low latitudes (<30°). The fatality was 6.76% in cold regions (<20°C) but only 3.90% in hot regions (≥20°C). The fatality was 5.87% in rainy regions (≥40mm) but only 3.33% in rainless regions (<40mm). The fatality was 6.57% in cloudy regions (<50) but only 3.86% in sunny regions (≥50). Traveling to hot sunny regions without pollution is a strategy for risk reduction.

## Introduction

The COVID-19 pandemic emerged in Wuhan in December 2019 and had led to 10 million confirmed cases and 0.5 million death by June 30, 2020.^1^ Impacts of regional weather and climate on the pandemic have been investigated widely. For example, one study revealed that daily temperature, humidity, wind speed, and UV index were associated with a lower incidence of COVID-19 by March 12.^2^ UV light was strongly associated with lower COVID-19 growth rates in the early phase before intervention.^3^ The temperature and humidity could explain 18% of the variation in case doubling time in China by March 1.^4^ Every 1°C increase in temperature was associated with a 3.08% reduction in daily new cases and a 1.19% reduction in daily new deaths in 166 countries by March 27.^5^ The doubling time of COVID-19 transmission increase by 40-50% when the temperature rised from 5°C to 25°C by April 14.^6^ The growth rate was affected by precipitation seasonality and warming velocity rather than temperature by April 6.^7^ A one-degree increase in absolute latitude was associated with a 2.6% increase in cases per million inhabitants.^8^

Experimental studies revealed that novel coronavirus could not be quickly destroyed by hot temperature but UV radiation in the summer. Ninety percent of infectious SARS-COV-2 virus was inactivated every 6.8 minutes in simulated saliva and every 14.3 minutes in culture media when exposed to simulated sunlight.^9^ However, UV light may be meaningless for inactivating viruses in areas with high air pollution where UV light turns into heat.^10^ COVID-19 exploded during the darkest January in Wuhan in over a decade, as daily irradiance correlated with case growth seven days later.^11^ The humid subtropical climate and serious pollution in Wuhan could partly explain why the case-fatality rate was up to 6.6% in Wuhan but only 0.8% in other regions of China.

## Methods

We investigated the climate impacts on COVID-19 fatality in 88 countries with more than 1000 confirmed cases and 100 deaths from December 2019 to June 2020 (table).

**Table.** Confirmed COVID-19 cases and deaths in 88 countries. *China includes Hong Kong, Macao, and Taiwan.

|  |  | Confirmed cases | Deaths | Case-fatality rate |
| --- | --- | --- | --- | --- |
| China* | 29 provinces and cities | 85021 | 4644 | 5.46% |
| United States | 53 state-level regions | 2652954 | 127453 | 4.80% |
| Other countries | 86 countries | 7331433 | 370424 | 5.05% |
| Total | 168 regions | 10069408 | 502521 | 4.99% |

We obtained province-level case data from the National Health Commission of China and its provincial branches.^12^ There were 29 provinces and cities with more than 100 confirmed cases in China. US state-level data was from The New York Times, based on reports from state and local health agencies.^13^ Every state-level epidemic region in 50 states, Washington DC, and two territories had more than 100 confirmed cases. We collected case data of each month from 168 epidemic regions across 88 countries. The number of monthly new cases and deaths were transformed by natural logarithm before analyses. These 168 regions are distributed in 12 climate regions based on Köppen climate classification: tropical rainforest, tropical grasslands, tropical monsoon, tropical desert, humid subtropical, Mediterranean climate, subtropical highland, subtropical desert, humid continental, temperate marine, temperate grasslands, temperate desert, subarctic.

We extracted latitude data from the Google map and climate data from Weather-atlas.^14^ We did not use the minimum temperature data but the maximum temperature in terms of collinearity. We calculated the means of absolute latitude, monthly maximum temperature, relative humidity, rainfall, and sunshine UV weighted by the number of monthly confirmed new cases. We created a new climate indicator, sunshine UV, the product of average sunshine time times average UV index. For example, the average sunshine is 7.5 hours per day and the average UV index is 6 in April, so the average sunshine UV is 45.

Correlation and nonlinear regression analyses were performed with Stata software to estimate the impacts of the climate on the number of monthly confirmed new cases and deaths. In order to evaluate the impacts of climate, all epidemic regions were divided into two latitude-groups, two temperature-groups, two humidity-groups, two rain-groups, and two UV-groups: high and low latitudes (≥30°, <30°), hot and cold regions (≥20°C, <20°C), humid and arid regions (≥60%, <60%), rainy regions and rainless regions (≥40mm/month, <40mm/month), sunny and cloudy regions (≥50/day, <50/day).

## Results

A total of 168 epidemic regions contributed 765 region-months in a mean follow-up of 4.55 months from December 2019 to June 2020. The seasonal pattern is inverted between the northern hemispheres (NH) and southern hemispheres (SH). Although the number of monthly confirmed new cases have continued to increase, there was a significant decrease in overall case-fatality rate in the NH in May and June as the summer was coming (from 8.30% to 4.98% and 3.17%). The number of monthly confirmed new deaths have continued to decrease in the NH but increase in the SH in May and June. There was a significant decrease in overall case-fatality rate in both the NH and SH in May and June (from 8.17% to 4.99% and 3.22%). Figure 1 shows monthly confirmed new cases and deaths of the COVID-19 pandemic in the NS and SH.

**Figure 1.**
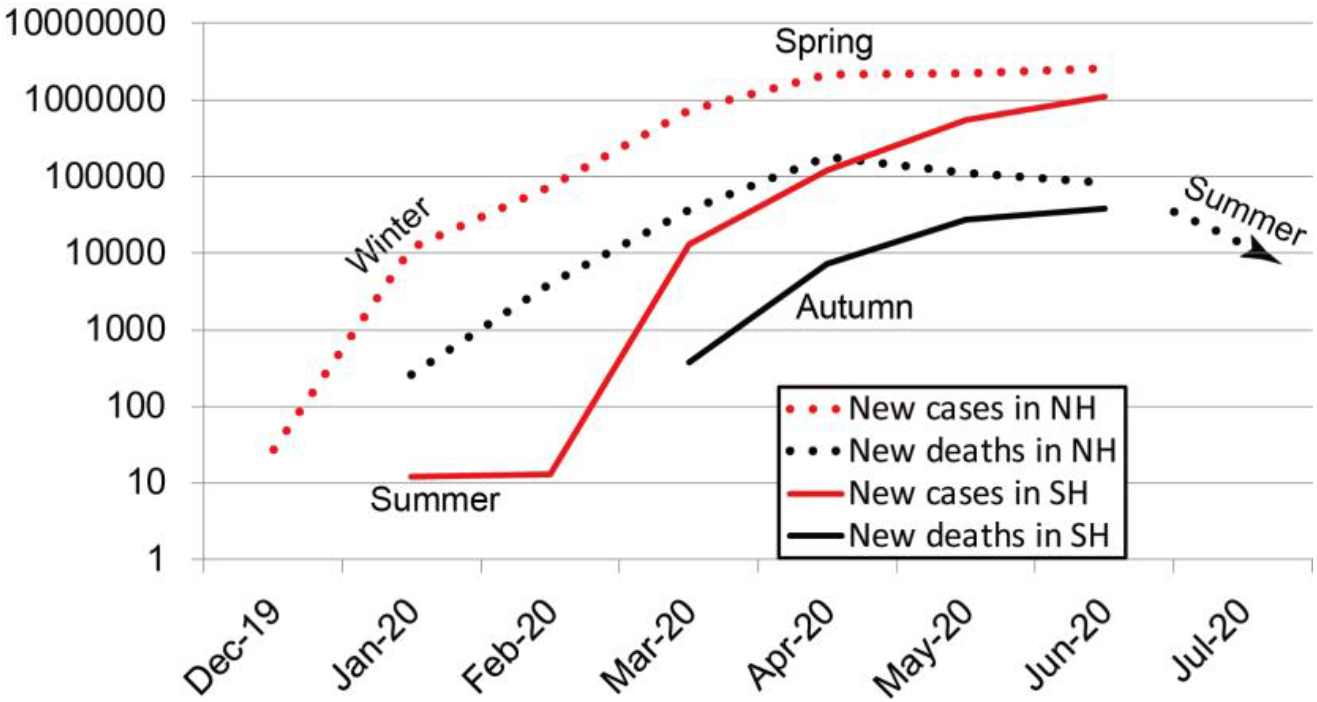
Monthly confirmed new cases and deaths in the NH and SH.

Although humid continental regions and humid subtropical regions had the most confirmed COVID-19 cases in all types of climate regions (20.15%, 20.06%), the case-fatality rates in the two regions were only 3.65% and 6.06%. The case-fatality rate in temperate marine climate regions was the highest (11.13%), including the United Kingdom, Ireland, France, Netherlands, Belgium, Denmark, Sweden, Norway, Germany, Poland, Czechia, Austria, Switzerland, Croatia, Luxembourg, and Slovenia. The case-fatality rate in tropical regions was only 3.20%, lower than subtropical regions and temperate regions (5.74%, 5.66%). Figure 2 shows the total number of confirmed cases and deaths of the COVID-19 pandemic in 12 climate regions.

**Figure 2.**
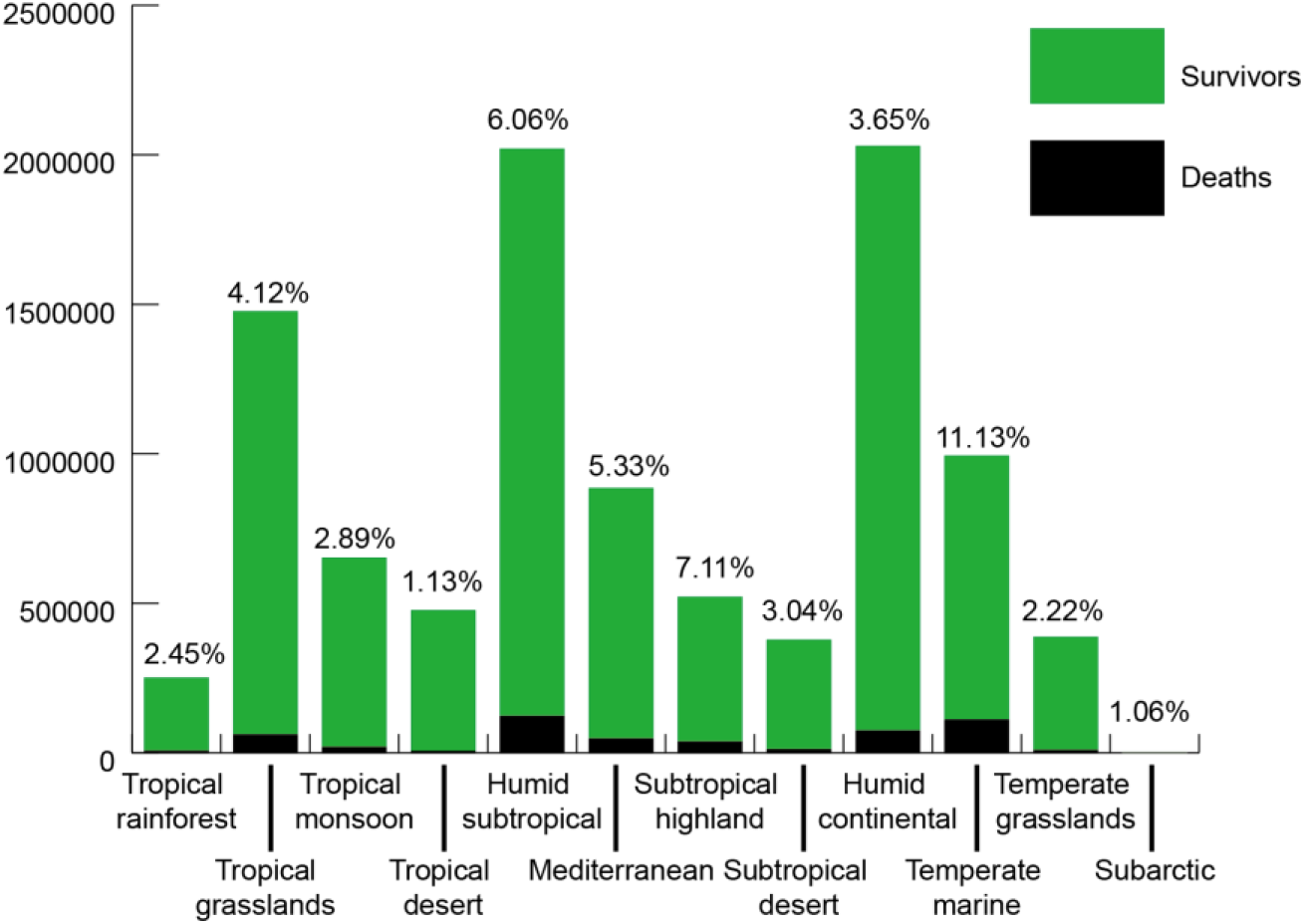
The COVID-19 fatality in different climate regions. The number of confirmed cases includes survivors (red bars) and deaths (black bars).

The means of absolute latitude, monthly maximum temperature, relative humidity, rainfall, and sunshine UV weighted by the number of monthly confirmed new cases are 32.9 degrees, 23.2°C, 62.8%, 61.8mm, and 61.3, respectively.

The number of confirmed cases in high latitudes (≥30°) was 62.3% of the total. The case-fatality rate was 5.71% in high latitudes but only 3.73% in low latitudes (<30°). Monthly average maximum temperature was strongly correlated to the sunshine UV (r=0.84). There was only a moderate correlation between relative humidity and rainfall (r=0.44). Figure 3 shows the COVID-19 fatality in different climate groups.

**Figure 3.**
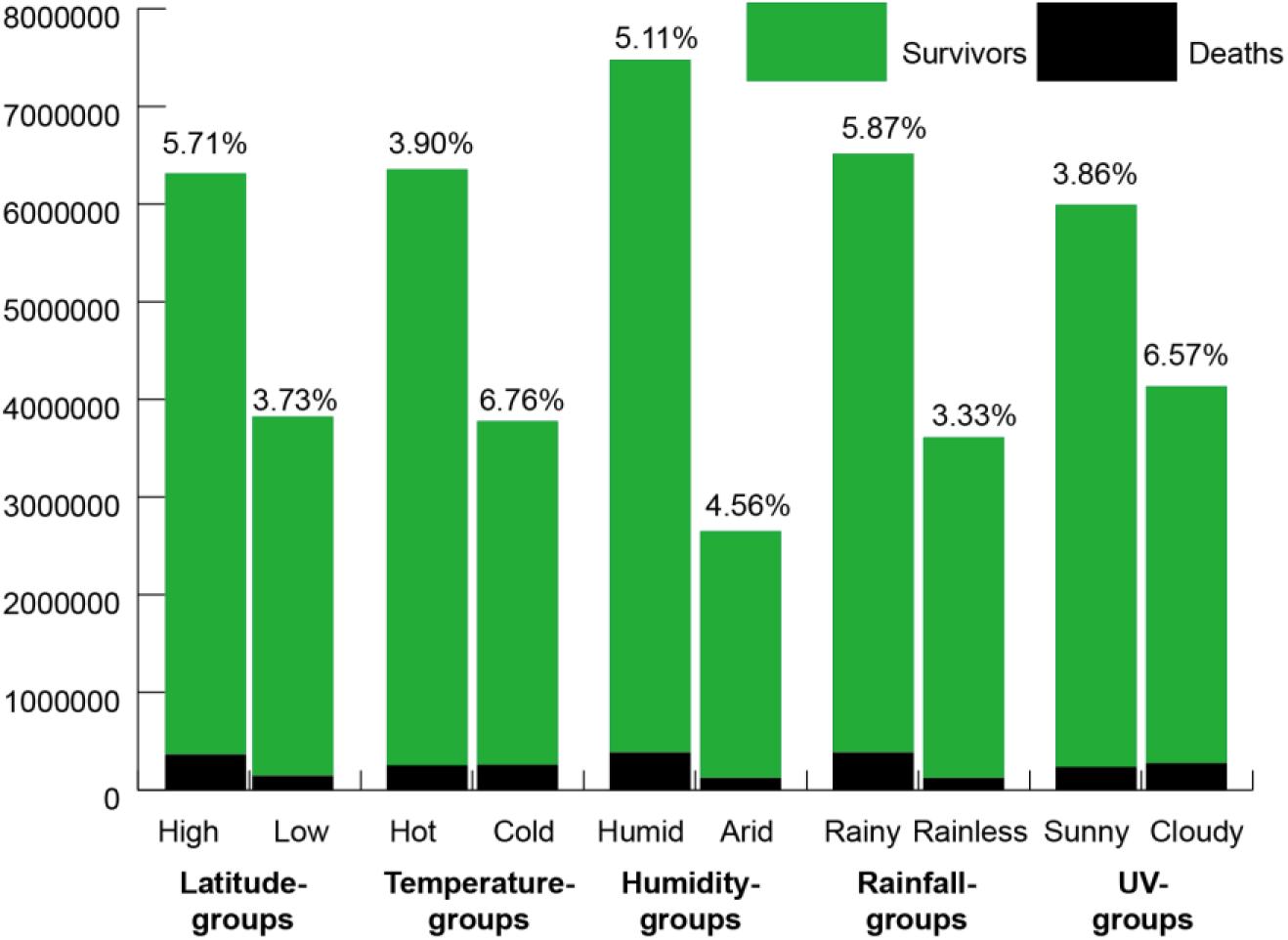
The COVID-19 fatality in different climate groups. The number of confirmed cases includes survivors (red bars) and deaths (black bars).

The number of confirmed cases in hot regions (≥20°C) was 62.7% of the total. The case-fatality rate was 6.76% in cold regions (<20°C) but only 3.90% in hot regions. The number of monthly confirmed cases and deaths increased as the temperature rised in cold regions (r=0.37, 0.38), but no correlation in hot regions.

A nonlinear regression analysis was performed to model the relationship between the number of monthly confirmed new cases (y) and monthly maximum temperature (x) in cold regions. The R-squared of this model is 0.87, *p*<0.0001. This regression model predicts that every 1°C increase in monthly maximum temperature leads to an increase in the natural logarithm of monthly confirmed new cases by 2.7% in cold regions. The model is as follows,

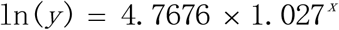

The number of confirmed cases in humid regions (≥60%) was 73.8% of the total. Monthly confirmed cases and deaths increased as the temperature rised in arid regions (r=0.35, 0.25), but only little correlation in humid regions (r=0.18, 0.18).

The number of confirmed cases in rainy regions (≥40mm/month) was 64.3% of the total. The case-fatality rate was 5.87% in rainy regions but only 3.33% in rainless regions (<40mm/month). Monthly confirmed new cases and deaths increased as the temperature rised in rainless regions (r=0.43, 0.39), but no correlation in rainy regions. Monthly confirmed new cases and deaths increased as the sunshine UV rised in rainless regions (r=0.44, 0.35), but no correlation in rainy regions.

The number of confirmed cases in sunny regions (UV≥50/day) was 59.2% of the total. The case-fatality rate was 6.57% in cloudy regions (UV<50/day) but only 3.86% in sunny regions (UV≥50/day). Monthly confirmed new cases and deaths increased as the temperature rised in cloudy regions (r=0.21, 0.34), but no correlation in sunny regions. Monthly confirmed new cases and deaths increased as the sunshine UV rised in cloudy regions (r=0.22, 0.30), but no correlation in sunny regions.

The case-fatality rate was 6.82% in cold humid regions but only 3.00% in hot arid regions. The case-fatality rate was 10.53% in cold cloudy regions but only 3.84% in hot sunny regions.

## Discussion

We found that the impacts of temperature, humidity, rainfall, and sunshine UV on the COVID-19 pandemic were consistent with several previous studies. Both SARS and COVID-19 emerged in the winter. SARS peaked in the spring and was gone in the summer of 2003. COVID-19 is considered to be the updated version of SARS, so we project the COVID-19 pandemic will show a downward trend in the NH in the coming summer.

Aerosol transmission of the virus by water microdroplets and pollution particles as carriers is plausible since the virus can remain viable and infectious in aerosols for hours and the UV radiation can destroy the virus in minutes. Many epicenters are humid and cloudy with air pollution and all kinds of viruses, forming so-called miasma. As the case-fatality rate was 12% in Northern Italy but only 4.5% in the rest of the country by March 21, 2020, researchers found a correlation between the high level of COVID-19 lethality and the atmospheric pollution in Northern Italy.^15^ An increase of only 1 μg/m^3^ in PM2.5 was associated with an 8% increase in the COVID-19 death rate in the United States up to April 22, 2020.^16^

It is wise to escape from an epicenter full of miasma. Traveling to hot sunny regions without pollution is a strategy for risk reduction. On the contrary, extremely locking down an epicenter to prevent escape is not the best policy. Wearing masks is necessary for epidemic prevention, especially in an epicenter full of miasma. Authorities should control air pollution, increasing fossil energy taxes to promote clean energy. UV lamps, air heaters, and oxygen bottles are necessary to improve the air quality in ICU and isolating rooms for patients. We suggest UV lamps should be compulsory in all service spaces, including classrooms, dormitories, hospitals, hotels, bars, restaurants, offices, warehouses, and public transport.

## Data Availability

All data needed to evaluate the conclusion in the paper are present in the paper or the supplementary materials.

## Contributors

MW proposed the project and wrote the paper. LC supported the project and revised the paper.

## Acknowledgments

We thank Deepak Gupta and Jessica Carr helpful feedback.

## Supplementary Materials

Figures S1-S4

Tables S1-S4

External database S1

